# Socio-demographic characteristics associated with COVID-19 vaccination uptake in Switzerland: longitudinal analysis of the CoMix study

**DOI:** 10.1101/2023.03.13.23287183

**Authors:** Martina L Reichmuth, Leonie Heron, Julien Riou, André Moser, Anthony Hauser, Nicola Low, Christian L Althaus

## Abstract

**Background:** Vaccination is an effective strategy to reduce morbidity and mortality from coronavirus disease 2019 (COVID-19). However, the uptake of COVID-19 vaccination has varied across and within countries. Switzerland has had lower levels of COVID-19 vaccination uptake in the general population than many other high-income countries. Understanding the socio-demographic factors associated with vaccination uptake can help to inform future vaccination strategies to increase uptake.

**Methods:** We conducted a longitudinal online survey in the Swiss population, consisting of six survey waves from June to September 2021. Participants provided information on socio-demographic characteristics, history of testing for severe acute respiratory syndrome coronavirus 2 (SARS-CoV-2), social contacts, willingness to be vaccinated, and vaccination status. We used a multivariable Poisson regression model to estimate the adjusted rate ratio (aRR) and 95% confidence intervals (CI) of COVID-19 vaccine uptake.

**Results:** We recorded 6,758 observations from 1,884 adults. For the regression analysis, we included 3,513 observations from 1,883 participants. By September 2021, 600 (75%) of 806 study participants had received at least one vaccine dose. Participants who were older, male, and students, had a higher educational level, household income, and number of social contacts, and lived in a household with a medically vulnerable person were more likely to have received at least one vaccine dose. Female participants, those who lived in rural areas and smaller households, and people who perceived COVID-19 measures as being too strict were less likely to be vaccinated. We found no significant association between previous SARS-CoV-2 infections and vaccination uptake.

**Conclusions:** Our results suggest that socio-demographic factors as well as individual behaviours and attitudes played an important role in COVID-19 vaccination uptake in Switzerland. Therefore, appropriate communication with the public is needed to ensure that public health interventions are accepted and implemented by the population. Tailored COVID-19 vaccination strategies in Switzerland that aim to improve uptake should target specific subgroups such as women, people from rural areas or people with lower socio-demographic status.

## Introduction

Vaccines can prevent symptomatic infections, severe disease, and death from coronavirus disease 2019 (COVID-19). The evidence of vaccine effectiveness comes from randomised clinical trials and real-world data (1,2). Although effective vaccines with a favourable safety profile are available against a wide range of pathogens, public confidence in vaccination has declined in some countries, and some population groups are increasingly reluctant to be vaccinated (3). The World Health Organization (WHO) ranks vaccine hesitancy among the top ten global health threats (4). Investigating the factors associated with vaccine hesitancy and lower vaccination uptake could help to develop strategies to minimise the impact of COVID-19 and future epidemics.

Several studies have reviewed factors that may be associated with COVID-19 vaccination uptake. A systematic review indicated that socio-demographic factors and perceptions of risk and susceptibility to COVID-19 were associated with the intention to get vaccinated and that vaccine attributes influenced vaccination intention, while receiving negative information about vaccines and working in healthcare resulted in lower intentions to get vaccinated (5). Switzerland has had lower levels of COVID-19 vaccine uptake in the general population than many other high-income countries (6). A prospective cohort study in Switzerland found that vaccination uptake was multifactorial and associated with socio-demographic characteristics, health status, trust in institutions, fears of side-effects and expected risk of severe COVID-19 (7). A further understanding of how socio-demographic and behavioural factors were associated with vaccine uptake, while accounting for the age-dependent roll-out during the COVID-19 vaccination program in Switzerland, will help to improve future vaccination strategies.

The objective of this study was to analyse the association of socio-demographic and other factors with COVID-19 vaccination uptake during the roll-out of the vaccination program in the general population in Switzerland. First, we conducted an online survey with six survey waves from June to September 2021. Second, we studied vaccination uptake in the survey population using a Poisson regression model. Finally, we investigated whether the participants’ characteristics were associated with missed survey waves.

## Methods

This study was conducted as part of the CoMix study, which is a longitudinal online survey about social contact patterns during the COVID-19 pandemic in more than 20 countries in Europe and is described in detail elsewhere (8,9). The questionnaire included socio-demographic characteristics, attitudes and practices towards public health interventions against COVID-19 and social contact behaviours. Questions about social contacts were based on the POLYMOD survey, conducted in 2008 (10).

In the longitudinal CoMix study design, a sample of the adult (≥18 years) Swiss population was invited by the market research company Ipsos MORI to take part in repeated survey waves. We aimed to include 1,000 participants per survey wave, who were representative of the population in Switzerland using quotas on age, gender, and region of residence. We compared the characteristics of the participants with Swiss demographic data as reported by the Federal Statistical Office (FSO) (11) and the vaccination uptake of the participants with the vaccination monitor from the Federal Office of Public Health (FOPH) (12). We used data from six online surveys from June to September 2021 (B1-B6). Enrolment of new participants continued over the first three waves, primarily due to inconsistent participation and to ensure a sufficient sample size.

Participants provided sociodemographic information, including age groups (categorised as 18-29, 30-39, 40-49, 50-59, 60-69, and 70+ years), gender (female or male), region (urban or rural), Swiss region of residence (nomenclature of territorial units for statistics (NUTS) regions of Switzerland), country of birth (Switzerland, European Union (EU), or non-EU), educational level (low (obligatory school and vocational education), middle (high school and advanced vocational education), and high (bachelor or higher)), employment level (unemployed, student, homemaker, retired, or unemployed due to health reasons), net household income (<5,000, 5,001-10,000, or >10,000 CHF, preferred not to answer), household size, and whether they were living in a household with a medically vulnerable individual, and testing for severe acute respiratory syndrome coronavirus 2 (SARS-CoV-2) (tested positive, tested, never tested, preferred not to answer). They also reported social contact behaviours (number of physical contacts per day), vaccination status, willingness to be vaccinated, and attitudes towards COVID-19 measures. Participation in the study was voluntary but each participant received 5 CHF per survey wave. We conducted all analyses using anonymised data in R version 4.2.1 and the code is available on GitHub: https://github.com/ISPMBern/comix. The study was approved by the ethics committee of the Canton of Bern (project number 2020-02926), all methods were performed in accordance with regulations, and informed consent of participants was obtained. We followed the STROBE Statement to report this study (13).

The primary outcome of the analysis was having received the first dose of the COVID-19 vaccine. In Switzerland, the first COVID-19 vaccine was approved in December 2020 (Swiss Agency for Therapeutic Products, Swissmedic 2020; Supplementary Table 1) and mRNA vaccines (Moderna and Pfizer-BioNTech) were most widely used. In addition, we reported the prevalence of fully vaccinated individuals in Switzerland by the end of our study period in September 2021 (defined as having received at least two doses).

We described vaccination uptake over time. First, we reported the willingness to be vaccinated as reported in the survey. Second, we modelled the primary outcome (vaccination uptake) as a point process using Poisson regression with the logarithm of the observation time (the length of the interval between follow-up surveys per participant, i.e., *t*_*i*_ -*t*_*i*-1_) as offset (or denominator) for vaccination uptake (14). Thus the unadjusted rates are given as follows: log(*y*_*i*_/(*t*_*i*_ -*t*_*i*-1_)) = *β*_0_+*β*x. We set time zero to be 1 January 2021, shortly after the administration of the first vaccinations. All participants’ observations were included until they reported having received the first dose, if applicable, and were censored thereafter. We included data recorded on unvaccinated participants at all timepoints. We derived rates from the exponentiated coefficients of the Poisson regression model.

Vaccination status was the dependent variable, and the following factors were covariates: time (survey wave), age, gender, region, residence, country of birth, education level, employment level, net household income, household size, vulnerable group within the household, testing for SARS-CoV-2, number of contacts, and attitude towards COVID-19 measures. The last three covariates could change over time for participants. We performed univariable and multivariable regression models and reported the rate ratio (RR) and adjusted RR (aRR) with 95% confidence intervals (CI), controlling for all covariates. We included time by survey waves and modelled an interaction with age to account for the different times at which vaccines became available for different age groups.

In a sensitivity analysis, we set time zero to be 1 June 2021, which was just before the first survey wave. We performed further sensitivity analyses and compared the results from the Poisson regression model to those derived using Cox proportional hazards regression models. We ran Cox regression models, first with all participants included in the main analysis and second for individuals with an exact date of vaccination (86%) or who had not been vaccinated during the study period, with and without inverse probability weighting cumulatively over time (IPWC) to account for dropouts (15). We estimated the probability of censoring with each observation or up to the time point of vaccination. Probabilities were then derived with a logistic regression model adjusting for all covariates as described in our main analysis. Further, we subtracted estimated probabilities from 1, which resulted in an estimate of the probability of being uncensored for a given observation. We accumulated these probabilities over time for each participant. Finally, we derived stabilised weights using a logistic regression model without covariates for the nominator. We defined missingness as when a participant was absent in any survey wave after recruitment. To estimate these probabilities, we used logistic regression with all observations and all covariates from the main regression model plus the primary outcome. Further, we use each participant’s last observation to test whether the missingness of a survey wave was associated with covariates that we previously described.

## Results

This study included six survey waves from 3 June 2021 to 9 September 2021, with participants enrolled during the first three waves (Table 1; Supplementary Figure 1; Supplementary Figure 2). We followed participants for 55 days on average (range: 0-103 days). The study included 6,758 observations from 1,884 participants. Overall, 918 (49%) were females and 956 (51%) were males. Participants’ age ranged from 18 to 90 years with a median of 47 years. The study population was largely representative of the Swiss population (Supplementary Table 2). For the regression analysis, we included 1,883 participants (one participant had missing data for vaccination status; Table 2). Further, we identified missing data for six observations from three participants (four for vaccination status and two for contact information). We excluded these observations from regression analyses. Of all who participated from June to September 2021, 443 (24%) did not miss any waves, 363 (19%) missed at least one survey wave, and 1,078 (57%) dropped out before the last wave.

**Table 1:** Overview of survey waves.

| Survey wave | Start date, year-month-day | End date, year-month-day | Number of participants | Number of newly enrolled participants | Number of missing participants who had been previously enrolled | Number of returning participants after missing at least one wave | Number of participants with no missing variables |
| --- | --- | --- | --- | --- | --- | --- | --- |
| B1 | 2021-06-03 | 2021-06-14 | 996 | 996 | 0 | 0 | 996 |
| B2 | 2021-07-02 | 2021-07-19 | 1,559 | 800 | 237 | 0 | 1,558 |
| B3 | 2021-07-20 | 2021-07-29 | 1,324 | 88 | 392 | 69 | 1,322 |
| B4 | 2021-08-10 | 2021-08-16 | 1,120 | 0 | 393 | 189 | 1,119 |
| B5 | 2021-08-26 | 2021-09-01 | 953 | 0 | 354 | 187 | 952 |
| B6 | 2021-09-09 | 2021-09-15 | 806 | 0 | 367 | 220 | 805 |

**Table 2:** Socio-demographic characteristics, history of testing for SARS-CoV-2, social contact behaviour, and perception of COVID-19 measures in all study participants and in study participants who got vaccinated by the end of the study. * For time-dependent variables, the last observation of the participant is given. Abbreviation: EU, European Union; CHF Swiss Francs; N, number of participants.

| Categories | All participants, N (%) | Vaccinated participants, N (%) |
| --- | --- | --- |
| **Total** | 1,883 (100%) | 1,321 (100%) |
| **Age groups, years** |  |  |
| 18-29 | 358 (19%) | 216 (16%) |
| 30-39 | 358 (19%) | 234 (18%) |
| 40-49 | 308 (16%) | 203 (15%) |
| 50-59 | 363 (19%) | 263 (20%) |
| 60-69 | 289 (15%) | 226 (17%) |
| 70+ | 207 (11%) | 179 (14%) |
| **Gender** |  |  |
| Female | 918 (49%) | 613 (46%) |
| Male | 955 (51%) | 699 (53%) |
| Other | 10 (1%) | 9 (1%) |
| **Region** |  |  |
| Urban | 1,426 (76%) | 1039 (79%) |
| Rural | 457 (24%) | 282 (21%) |
| **Swiss regions** |  |  |
| Espace Mittelland | 406 (22%) | 273 (21%) |
| Zurich | 351 (19%) | 260 (20%) |
| Lake Geneva region | 337 (18%) | 235 (18%) |
| Eastern Switzerland | 263 (14%) | 186 (14%) |
| Northwestern Switzerland | 262 (14%) | 181 (14%) |
| Central Switzerland | 182 (10%) | 129 (10%) |
| Ticino | 82 (4%) | 57 (4%) |
| **Country of birth** |  |  |
| Switzerland | 1,331 (71%) | 927 (70%) |
| EU | 249 (13%) | 176 (13%) |
| Non-EU | 156 (8%) | 113 (9%) |
| Unknown | 147 (8%) | 105 (8%) |
| **Education level** |  |  |
| Obligatory school and vocational education | 805 (43%) | 531 (40%) |
| Gymnasium and advanced vocational education | 639 (34%) | 439 (33%) |
| Higher education (e.g., Bachelor, Master, or PhD) | 439 (23%) | 351 (27%) |
| **Employment status** |  |  |
| Employed | 1,161 (62%) | 789 (60%) |
| Unemployed | 110 (6%) | 67 (5%) |
| Student | 116 (6%) | 83 (6%) |
| Homemaker | 75 (4%) | 43 (3%) |
| Retired | 377 (20%) | 313 (24%) |
| Another unemployed situation | 44 (2%) | 26 (2%) |
| **Household income, net** |  |  |
| 0-5,000 CHF | 592 (31%) | 380 (29%) |
| 5,001-10,000 CHF | 762 (40%) | 539 (41%) |
| 10,000+ CHF | 248 (13%) | 200 (15%) |
| Preferred not to answer | 281 (15%) | 202 (15%) |
| **Household size** |  |  |
| Median (range) | 2 (1-10) | 2 (1-10) |
| **Household with medically vulnerability** |  |  |
| No person in a risk group | 1,305 (69%) | 871 (66%) |
| One or more person in a risk group | 578 (31%) | 450 (34%) |
| **Testing for SARS-Cov-2\*** |  |  |
| Tested positive | 31 (2%) | 25 (2%) |
| Tested | 543 (29%) | 336 (25%) |
| Never tested | 1,277 (68%) | 941 (71%) |
| Preferred not to answer | 32 (2%) | 19 (1%) |
| **Number of contacts per day\*** |  |  |
| 0-2 | 767 (41%) | 547 (41%) |
| 3-5 | 527 (28%) | 377 (29%) |
| 6+ | 589 (31%) | 397 (30%) |
| **Attitudes towards COVID-19 measures\*** |  |  |
| About right | 913 (48%) | 737 (56%) |
| Too lenient | 423 (22%) | 356 (27%) |
| Too strict | 501 (27%) | 204 (15%) |
| Don't know | 46 (2%) | 24 (2%) |

From May 2021 onwards, the COVID-19 vaccination campaign in Switzerland targeted the entire adult population and uptake increased during the study period (Figure 1A). Vaccination uptake in our study population was higher than in the overall population of Switzerland. In the first survey wave of June 2021, 533 (54%) had at least one vaccine dose compared with 43% of the general Swiss population. This increased to 75% by the sixth survey wave, compared with 70% in the general adult population (Figure 1A). Participants who had not already been vaccinated indicated their willingness as whether they intended, were hesitating, or had no intention to get vaccinated. The increase in vaccine uptake within the CoMix study occurred mainly amongst those who wanted to get vaccinated (18% in the first wave to 4% in the last wave) rather than those that had no intention (16% in the first wave to 14% in the last wave) or were hesitant (12% in the first wave to 7% in the last wave) (Figure 1B).

**Figure 1:**
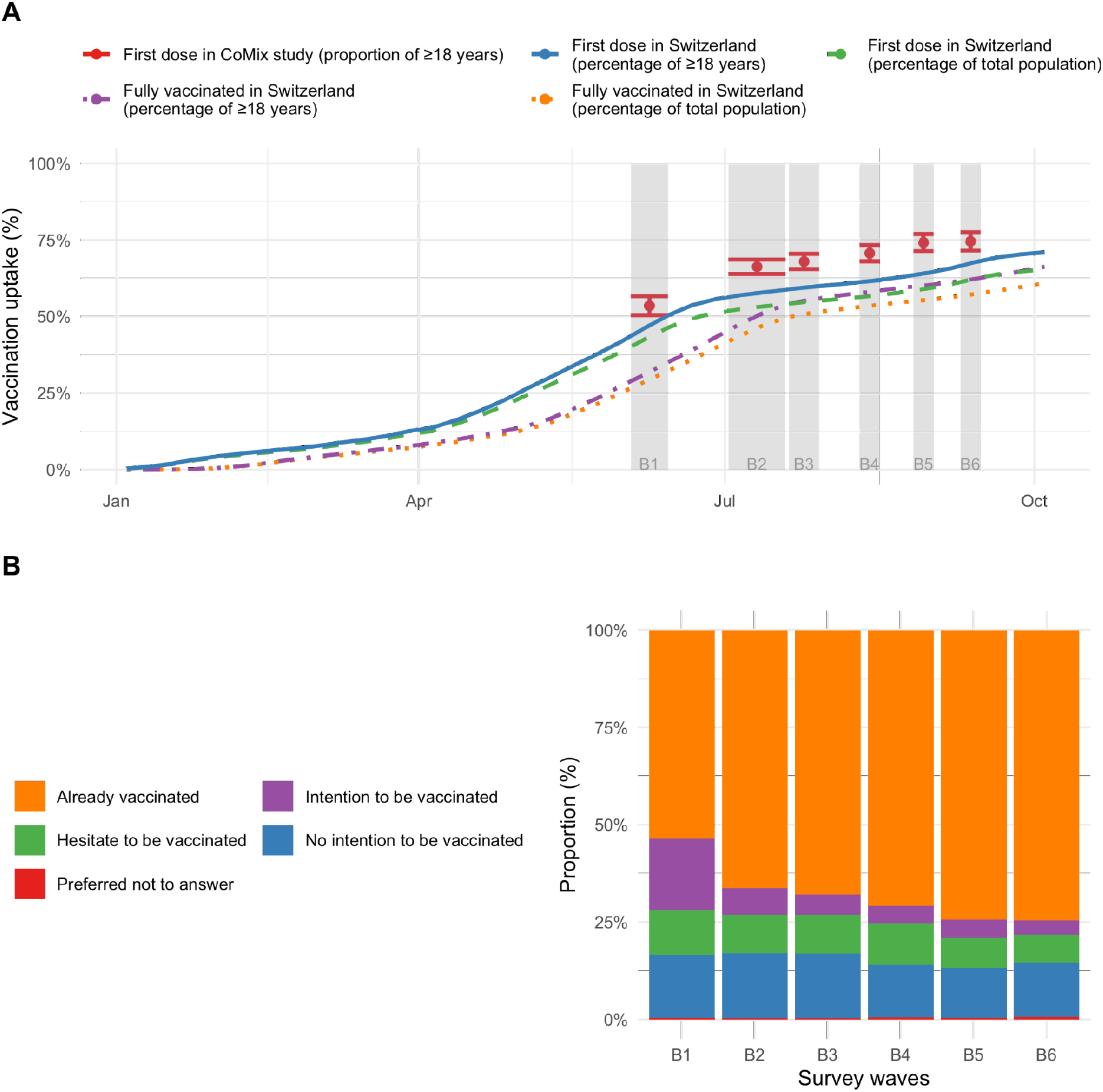
COVID-19 vaccination uptake in Switzerland. A: Comparison of vaccination uptake in the CoMix survey participants (red dots with 95% confidence intervals) and general population of Switzerland. B: Willingness to receive COVID-19 vaccination.

In the Poisson regression model, we found that people in all older age groups were more likely to get vaccinated than those in the youngest age group (18-29 years; Figure 2; Figure 3; Supplementary Figure 3). In adults 30 years and older, vaccination uptake per interval was highest before the first survey wave and declined afterwards. The vaccination uptake per interval in younger adults (18-29 years) peaked at the second survey wave, then declined and increased again at the last survey wave. Being male was associated with higher vaccination uptake (aRR 1.09, 95% CI: 1.04-1.15) (Figure 2). We found geographical differences in vaccination uptake. Living in rural areas was associated with lower vaccine uptake than in urban areas (aRR 0.85, 95% CI: 0.80-0.90). Vaccination uptake varied slightly between regions. Most regions were associated with higher vaccine uptake than Espace Mittelland. We did not find statistical evidence of an association between country of birth and vaccination uptake.

**Figure 2.**
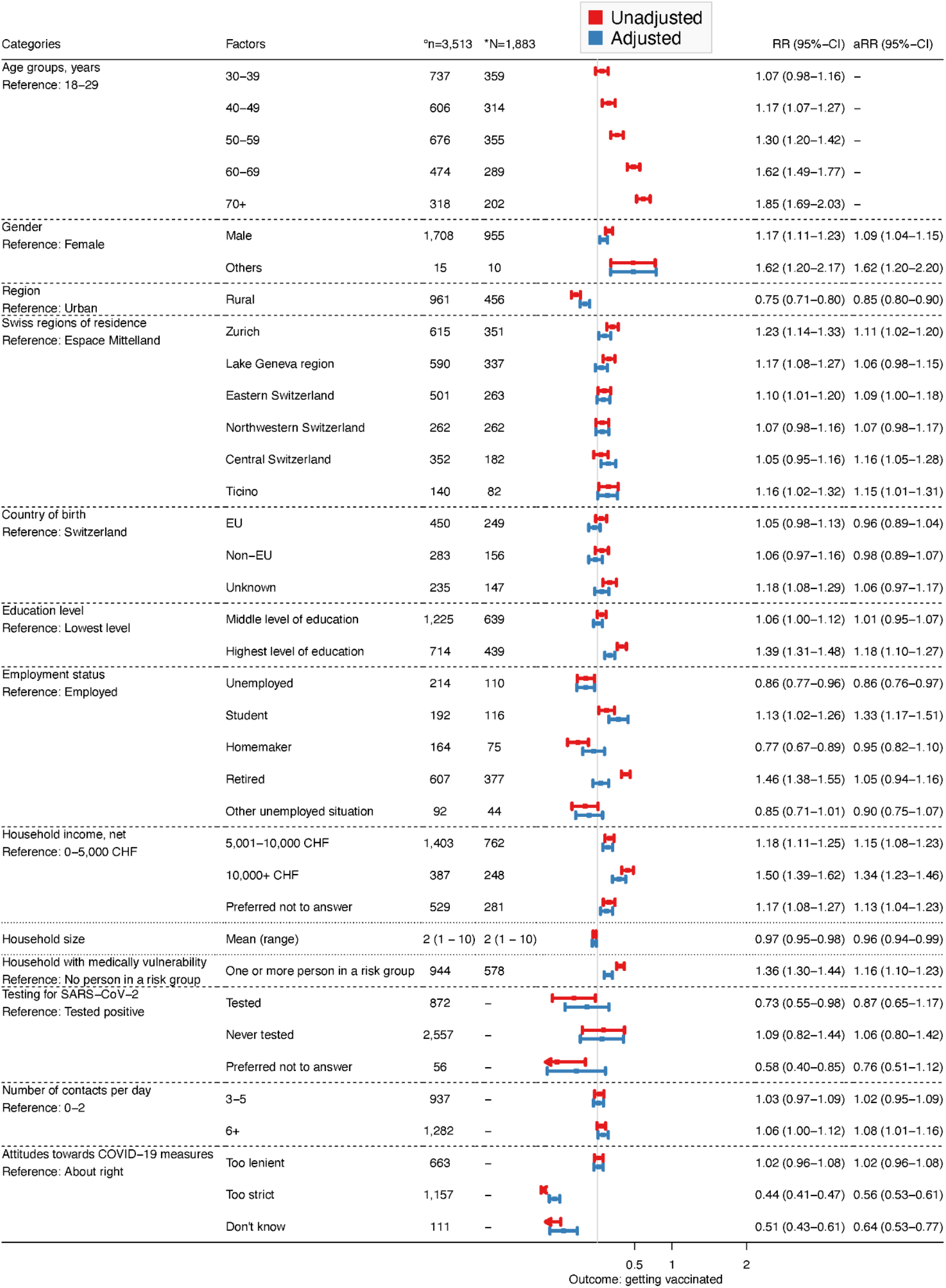
Results of the univariable and multivariable Poisson regression models. The primary outcome of the analysis is having received the first dose of a COVID-19 vaccine. °Number (n) of observations included in the regression analysis. *Number (N) of participants included in the regression analysis. Abbreviations: EU, European Union; CHF, Swiss Francs; CI, confidence interval; RR, rate ratio; aRR, adjusted rate ratio.

**Figure 3.**
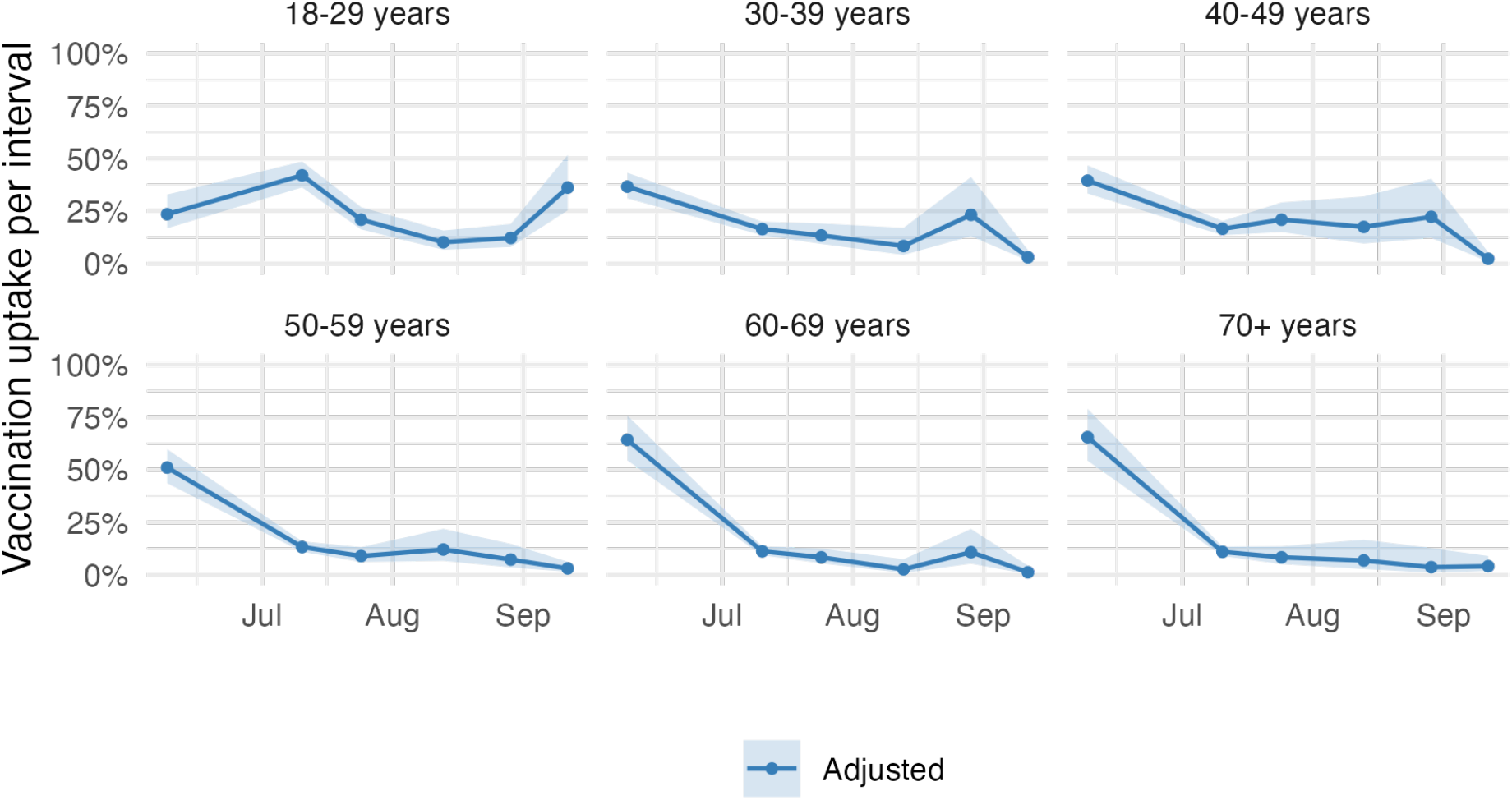
Vaccination uptake per interval by age group. Vaccination uptake corresponds to the percentage receiving the first vaccine dose amongst those who have not already received it. The adjusted estimates were adjusted for age with an interaction with survey wave, gender, region, Swiss region of residence, country of birth, education level, employment level, net household income, household size, household with a medically vulnerable individual, testing for SARS-CoV-2, number of contacts per day, and attitude towards COVID-19 measures. The shaded area indicates the 95% confidence interval.

We found that the highest education level (having a Bachelor, Master or PhD), was associated with a higher vaccination uptake (aRR 1.18, 95% CI: 1.10-1.27) than with the lowest education level (completed obligatory school and vocational education only). Unemployed participants were less likely (aRR 0.86, 95% CI: 0.76-0.97) and students were more likely (aRR: 1.33, 95% CI: 1.17-1.51) to get vaccinated than employed participants. In addition, higher income was associated with higher vaccination uptake. A household income between 5,001 CHF and 10,000 CHF compared with less than 5,000 CHF resulted in an aRR of 1.15 (95% CI: 1.08-1.23) and an income of at least 10,000 CHF resulted in an aRR of 1.34(95% CI: 1.23-1.46) Living in smaller households was associated with lower vaccination uptake (aRR 0.96, 95% CI: 0.94-0.99). In contrast, living with a medically vulnerable individual was associated with a higher aRR of 1.16 (95% CI: 1.10-1.23). We found no association between previous SARS-CoV-2 infections and vaccination uptake. Individuals with six or more contacts per day had higher vaccination uptake than those with fewer than three contacts (aRR 1.08, 95% CI: 1.01-1.16). We also found that the perception of COVID-19 measures was associated with vaccination uptake. Participants who thought that the control measures were too strict were less likely to be vaccinated compared to those who thought that the control measures were about right (aRR 0.56, 95% CI: 0.53-0.61).

Setting time zero to 1 June 2021, did not substantially change the results of the Poisson regression model (Supplementary Table 3). The results from the Cox regression model were similar compared to those from the Poisson regression model (Supplementary Table 3 and 4). However, we deemed the Cox regression model less appropriate for the analysis of the data because the strong correlation between age and the time point of vaccination as a result of the age-specific vaccination campaign violates the proportional hazard assumption.

We also studied whether certain characteristics of participants were associated with missed survey waves (n=1,441, 76%). We found that individuals between 40 and 69 years were less likely to have missed survey waves than the youngest age group (18-29 years). Participants living in Geneva and Ticino were more likely to have missed survey waves compared to those living in Espace Mittelland. The same was found for those born in an EU country than those born in Switzerland. Participants with six or more contacts were also more likely to have missed survey waves than those with fewer than three contacts. We did not find strong statistical evidence for associations between missingness and gender, region, education level, employment status, household income, history of testing for SARS-CoV-2, or vaccination (Supplementary Table 5). Participants who missed survey waves had little impact on the results from the Cox regression model, as the unweighted and weighted hazard ratios (HR) were similar (Supplementary Table 3).

## Discussion

This study presents findings from analyses investigating factors associated with COVID-19 vaccination uptake in participants in the CoMix study in Switzerland. We found that vaccination uptake differed between subgroups from June to September 2021, a period during which COVID-19 vaccines were available to the entire adult population in Switzerland. Individuals who were older, male, and students, had a higher educational level, household income, and number of social contacts, and lived in a household with a medically vulnerable person were associated with higher vaccination uptake. In contrast, individuals who lived in rural areas, smaller households, and who perceived COVID-19 measures too strict were associated with lower uptake. There was no significant association between previous SARS-CoV-2 infections and vaccination uptake. Together, these results suggest that socio-demographic factors as well as individual behaviour and attitudes shaped COVID-19 vaccination uptake in Switzerland.

A major strength of our study is the use of the longitudinal CoMix survey to study multiple factors that are associated with COVID-19 vaccination uptake. The survey was based on quotas on age, gender, and region of residence and aimed to be representative of the Swiss population. As a result of the longitudinal data collection over six survey waves and modelling vaccination uptake as a point process using a Poisson regression model, we were able to capture changes in social contacts and attitudes on control measures over time. In France, Germany, and Italy, the introduction of COVID-19 vaccine passports in September 2021 resulted in an increase in vaccination uptake (16). In our study, the increase in vaccination uptake in 18-49 year olds during the last two survey waves at the end of summer 2021 coincided with the introduction of a COVID-19 vaccination certificate, which was required for participation in certain activities (Supplementary Table 1). Another possible reason for the increase in the vaccination uptake in late summer 2021 could be the easier scheduling of vaccination appointments after the summer holidays. In contrast to the study by Heininger et al. (7), we were also able to study the association of previous SARS-CoV-2 infections and the number of social contacts with COVID-19 vaccination uptake in Switzerland.

Our study also comes with a number of limitations. The potential inclusion of participants from the same household may influence our findings due to shared behaviours among household members. However, we think that such an event would be highly unlikely due to the survey design and the recruitment of participants. Further, the overall vaccination uptake in the study population by September 2021 (75%) was somewhat higher compared to the Swiss adult population (70%). This difference could be a result of the recruitment method within which the CoMix study was biased towards individuals with access to the internet, who may be reached by banner ads, email campaigns, and social media advertisements. In addition, survey participants are likely to be healthier than the general population (17). In the context of the CoMix study, participants might be more health-conscious and more likely to be vaccinated than the general population. Moreover, we found that individuals from the youngest and oldest age groups, non-German speaking regions, who were born in an EU country, and who had a higher number of contacts were more likely to have missed a survey wave. Therefore, the vaccination uptake and the aRR for these categories could be slightly underestimated. Although accounting for missing data from participants who missed survey waves hardly affected estimated HRs, associations between the place of residence, place of birth, and contact number with vaccination uptake should be interpreted with caution. As indicated by Moser et al. (2018), relative outcome measures like RRs may be less prone to bias than absolute quantities (18). Further, we did not collect information about the political orientation of participants, which may have an association with COVID-19 vaccination uptake as found for the United States but not for the United Kingdom (19,20).

Our analysis indicated that older age and higher socio-demographic status were associated with higher COVID-19 vaccination uptake in Switzerland, similar to the findings of some other studies (7,19,21–23). Lazarus et al. have, however, observed considerable heterogeneity in vaccine acceptance between countries (24). Vaccine hesitancy has also been shown to vary substantially at county level within the US (25). For example, gender as a predictor of COVID-19 vaccine acceptance and hesitancy varied globally (7,24,26,27). In our study, women reported lower vaccination uptake than men, possibly due to the mixed guidance for pregnant women or women wanting to become pregnant (28,29). Among women, Skjefte et al. also found that younger age, lower income, lower level of education, being unmarried and not having health insurance were associated with vaccine hesitancy (30). We did not find a significant association between place of birth and vaccination uptake, but systematic reviews indicated low intent to get vaccinated and low uptake in some migrant population groups (31,32). We asked participants’ about their perception of current COVID-19 measures, which might reflect trust in the government, which was found to be decisive in vaccine uptake (26). Moreover, Lazarus et al. stated that vaccine hesitancy is associated with a lack of trust in COVID-19 vaccine safety and science, and scepticism about vaccine efficacy (24). Finally, we found that individuals with a higher daily number of social contacts had a higher vaccination uptake. This could either be a result of participants increasing their number of contacts after vaccination, or that participants with a higher number of contacts are more willing to get vaccinated to protect themselves and others from infection, severe disease, and death.

Decision-making about vaccination strategies often occurs in the presence of uncertainties (25). To develop tailored and effective vaccination strategies, it is important to understand the multifactorial causes and context of vaccination hesitancy (22). Factors associated with vaccine hesitancy or uptake, often encompass political, religious, and socioeconomic aspects, but might vary across time, location, and specific vaccines (33,34). Despite the difference in time and context, a study examining the uptake of human papillomavirus (HPV) vaccines in Switzerland also found that individuals living in rural areas tended to be vaccinated less frequently (35). Vaccination strategies need to be carefully planned to ensure readiness of both the public and the health community, including the need for effective communication about the complexities of vaccination, such as the recognition that side-effects may occur shortly after vaccination while protection from severe disease only follows later. Vaccination strategies also require a broad range of approaches on the individual, provider, health system, and national levels, which is difficult to properly coordinate and promote (36). Policymakers have historically considered multiple options to increase vaccine uptake, ranging from communication and outreach strategies to monetary (dis)incentives, encouraging parental responsibility, and minimising distrust of expertise (37). Experts, such as physicians and other health care providers, are still among the most trusted individuals when it comes to health care advice, including for vaccination (20,25,38). Both, science, and health professionals, should be adequately trained in knowledge communication. Low vaccine uptake might be due to access and communication barriers and highlight that it is key to have outreach, and credible, consistent, and unified information about vaccines (3), such as that vaccines are among the most effective measures ever achieved through medical intervention. Engaging with and comprehending individuals sceptical about vaccination is of importance. In our study, we observed minimal changes in the attitudes of individuals who expressed no intention to get vaccinated (16% vs. 14% maintained their attitude throughout the study). We showed, within another panel of participants in the CoMix study in Switzerland, that almost half of individuals who did not intend to be vaccinated lacked trust in vaccines or feared side effects (39). Horne et al. (2015) underscored the positive influence of factual information on people’s attitudes towards vaccination in relation to communicable disease risks (40). Future research should focus on exploring effective social intervention strategies to enhance the uptake of vaccination. Finally, transparency about vaccine effectiveness and adverse events to set public expectations should improve trust in vaccines, but messaging should take care to avoid unintentionally overemphasising the risk of rare adverse events (41).

Our analysis suggests that women and individuals from rural areas, people with lower levels of education and lower household income, those who were unemployed, and who perceived the pandemic measures as being too strict were less likely to get vaccinated against COVID-19 in Switzerland. Tailored vaccination strategies towards these communities with lower vaccination uptake can be decisive as COVID-19 vaccination remains an important pillar in preventing severe disease and death.

## Data Availability

Scripts used for the analysis are available on GitHub: https://github.com/ISPMBern/comix.

## Declarations

### Ethics approval and consent to participate

The CoMix study protocols and questionnaires were approved by the local ethics committee of the Canton of Bern (project number 2020-02926), all methods were performed in accordance with regulations, and informed consent of participants was obtained.

### Consent for publication

Not applicable.

### Availability of data and materials

Scripts used for the analysis are available on GitHub: https://github.com/ISPMBern/comix

### Author contributions

MR, LH, AM, NL, and CA conceived and designed the study. MR performed the analysis and wrote the first draft. MR, LH, AM, NL, JR, AH, and CA contributed to the interpretation of the results. MR, LH and CA wrote the manuscript. All authors commented on the manuscript and approved the final version.

## Acknowledgements

We like to thank the European Centre for Disease Prevention and Control (ECDC) and the CoMix Europe Working Group for setting up the CoMix study across more than 20 European countries, and the partners at Ipsos MORI for running the survey.

## References

1. Pouwels KB, Pritchard E, Matthews PC, Stoesser N, Eyre DW, Vihta KD, et al. Effect of Delta variant on viral burden and vaccine effectiveness against new SARS-CoV-2 infections in the UK. Nat Med. 2021 Dec;27(12):2127–35.

2. Dickerman BA, Madenci AL, Gerlovin H, Kurgansky KE, Wise JK, Figueroa Muñiz MJ, et al. Comparative Safety of BNT162b2 and mRNA-1273 Vaccines in a Nationwide Cohort of US Veterans. JAMA Intern Med. 2022 Jul 1;182(7):739.

3. Black S, Rappuoli R. A Crisis of Public Confidence in Vaccines. Sci Transl Med [Internet]. 2010 Dec 8 [cited 2022 Sep 13];2(61). Available from: https://www.science.org/doi/10.1126/scitranslmed.3001738

4. World Health Organization (WHO). Ten threats to global health in 2019 [Internet]. 2019 [cited 2022 Jul 15]. Available from: https://www.who.int/news-room/spotlight/ten-threats-to-global-health-in-2019

5. Al-Amer R, Maneze D, Everett B, Montayre J, Villarosa AR, Dwekat E, et al. COVID-19 vaccination intention in the first year of the pandemic: A systematic review. J Clin Nurs. 2022 Jan;31(1–2):62–86.

6. Ritchie H, Edouard M, Lucas RG, Appel C, Giattino C, Ortiz-Ospina E, et al. Coronavirus Pandemic (COVID-19). Our World Data [Internet]. 2020; Available from: https://ourworldindata.org/coronavirus

7. Heiniger S, Schliek M, Moser A, von Wyl V, Höglinger M. Differences in COVID-19 vaccination uptake in the first 12 months of vaccine availability in Switzerland – a prospective cohort study. Swiss Med Wkly [Internet]. 2022 Mar 28 [cited 2022 Sep 12];152(13–14). Available from: https://smw.ch/article/doi/smw.2022.w30162

8. Jarvis CI, Van Zandvoort K, CMMID COVID-19 working group, Gimma A, Prem K, Klepac P, et al. Quantifying the impact of physical distance measures on the transmission of COVID-19 in the UK. BMC Med. 2020 Dec;18(1):124.

9. Verelst F, Hermans L, Vercruysse S, Gimma A, Coletti P, Backer JA, et al. SOCRATES-CoMix: a platform for timely and open-source contact mixing data during and in between COVID-19 surges and interventions in over 20 European countries. BMC Med. 2021 Dec;19(1):254.

10. Mossong J, Hens N, Jit M, Beutels P, Auranen K, Mikolajczyk R, et al. Social Contacts and Mixing Patterns Relevant to the Spread of Infectious Diseases. Riley S, editor. PLoS Med. 2008 Mar 25;5(3):e74.

11. Federal Statistical Office. Population [Internet]. [cited 2023 Jan 26]. Available from: https://www.bfs.admin.ch/bfs/en/home/statistics/population.html

12. Swiss Federal Office Public Health (FOPH). COVID-19 Switzerland Dashboard [Internet]. [cited 2021 Sep 14]. Available from: https://www.covid19.admin.ch/

13. Vandenbroucke JP. Strengthening the Reporting of Observational Studies in Epidemiology (STROBE): Explanation and Elaboration. Ann Intern Med. 2007 Oct 16;147(8):W.

14. Aalen OO, Borgan Ø, Gjessing HK. Survival and Event History Analysis [Internet]. New York, NY: Springer New York; 2008 [cited 2023 Feb 9]. (Gail M, Krickeberg K, Samet J, Tsiatis A, Wong W, editors. Statistics for Biology and Health). Available from: http://link.springer.com/10.1007/978-0-387-68560-1

15. Fewell Z, Hernán MA, Wolfe F, Tilling K, Choi H, Sterne JAC. Controlling for Time-dependent Confounding using Marginal Structural Models. Stata J Promot Commun Stat Stata. 2004 Dec;4(4):402–20.

16. Oliu-Barton M, Pradelski BSR, Woloszko N, Guetta-Jeanrenaud L, Aghion P, Artus P, et al. The effect of COVID certificates on vaccine uptake, health outcomes, and the economy. Nat Commun. 2022 Dec;13(1):3942.

17. Keyes KM, Rutherford C, Popham F, Martins SS, Gray L. How Healthy Are Survey Respondents Compared with the General Population?: Using Survey-linked Death Records to Compare Mortality Outcomes. Epidemiology. 2018 Mar;29(2):299–307.

18. Moser A, Bopp M, Zwahlen M, Swiss National Cohort study group. Calibration adjustments to address bias in mortality analyses due to informative sampling—a census-linked survey analysis in Switzerland. PeerJ. 2018 Feb 13;6:e4376.

19. Klymak M, Vlandas T. Partisanship and Covid-19 vaccination in the UK. Sci Rep. 2022 Nov 18;12(1):19785.

20. Albrecht D. Vaccination, politics and COVID-19 impacts. BMC Public Health. 2022 Dec;22(1):96.

21. Terry E, Cartledge S, Damery S, Greenfield S. Factors associated with COVID-19 vaccine intentions during the COVID-19 pandemic; a systematic review and meta-analysis of cross-sectional studies. BMC Public Health. 2022 Sep 2;22(1):1667.

22. Dubé E, MacDonald NE. COVID-19 vaccine hesitancy. Nat Rev Nephrol. 2022 Jul;18(7):409–10.

23. Leos-Toro C, Ribeaud D, Bechtiger L, Steinhoff A, Nivette A, Murray AL, et al. Attitudes Toward COVID-19 Vaccination Among Young Adults in Zurich, Switzerland, September 2020. Int J Public Health. 2021 May 6;66:643486.

24. Lazarus JV, Ratzan SC, Palayew A, Gostin LO, Larson HJ, Rabin K, et al. A global survey of potential acceptance of a COVID-19 vaccine. Nat Med. 2021 Feb;27(2):225–8.

25. Larson HJ, Gakidou E, Murray CJL. The Vaccine-Hesitant Moment. Longo DL, editor. N Engl J Med. 2022 Jul 7;387(1):58–65.

26. de Figueiredo A, Larson HJ. Exploratory study of the global intent to accept COVID-19 vaccinations. Commun Med. 2021 Sep 9;1(1):30.

27. Detoc M, Bruel S, Frappe P, Tardy B, Botelho-Nevers E, Gagneux-Brunon A. Intention to participate in a COVID-19 vaccine clinical trial and to get vaccinated against COVID-19 in France during the pandemic. Vaccine. 2020 Oct;38(45):7002–6.

28. Wong KLM, Gimma A, Paixao ES, CoMix Europe Working Group, Paolotti D, Karch A, et al. Pregnancy during COVID-19: social contact patterns and vaccine coverage of pregnant women from CoMix in 19 European countries. BMC Pregnancy Childbirth. 2022 Oct 8;22(1):757.

29. Stock SJ, Carruthers J, Calvert C, Denny C, Donaghy J, Goulding A, et al. SARS-CoV-2 infection and COVID-19 vaccination rates in pregnant women in Scotland. Nat Med. 2022 Mar;28(3):504–12.

30. Skjefte M, Ngirbabul M, Akeju O, Escudero D, Hernandez-Diaz S, Wyszynski DF, et al. COVID-19 vaccine acceptance among pregnant women and mothers of young children: results of a survey in 16 countries. Eur J Epidemiol. 2021 Feb;36(2):197–211.

31. Dubé E, Gagnon D, Nickels E, Jeram S, Schuster M. Mapping vaccine hesitancy—Country-specific characteristics of a global phenomenon. Vaccine. 2014 Nov;32(49):6649–54.

32. Crawshaw AF, Farah Y, Deal A, Rustage K, Hayward SE, Carter J, et al. Defining the determinants of vaccine uptake and undervaccination in migrant populations in Europe to improve routine and COVID-19 vaccine uptake: a systematic review. Lancet Infect Dis. 2022 Sep;22(9):e254–66.

33. Larson HJ, Jarrett C, Eckersberger E, Smith DMD, Paterson P. Understanding vaccine hesitancy around vaccines and vaccination from a global perspective: A systematic review of published literature, 2007–2012. Vaccine. 2014 Apr;32(19):2150–9.

34. MacDonald NE. Vaccine hesitancy: Definition, scope and determinants. Vaccine. 2015 Aug;33(34):4161–4.

35. Riesen M, Konstantinoudis G, Lang P, Low N, Hatz C, Maeusezahl M, et al. Exploring variation in human papillomavirus vaccination uptake in Switzerland: a multilevel spatial analysis of a national vaccination coverage survey. BMJ Open. 2018 May;8(5):e021006.

36. McIntosh EDG, Janda J, Ehrich JHH, Pettoello-Mantovani M, Somekh E. Vaccine Hesitancy and Refusal. J Pediatr. 2016 Aug;175:248–249.e1.

37. WHO SAGE working group dealing with vaccine hesitancy. Strategies for addressing vaccine hesitancy– A systematic review [Internet]. Available from: https://cdn.who.int/media/docs/default-source/immunization/sage/2014/october/3-sage-wg-strategies-addressing-vaccine-hesitancy-2014.pdf?sfvrsn=b632b81e_4

38. Featherstone JD, Bell RA, Ruiz JB. Relationship of people’s sources of health information and political ideology with acceptance of conspiratorial beliefs about vaccines. Vaccine. 2019 May;37(23):2993–7.

39. Reichmuth ML, Heron L, Low N, Althaus CL. Social contacts and attitudes towards vaccination during the COVID-19 pandemic: Insights from the CoMix study. [Internet]. 2022 Sep [cited 2023 May 27] p. Bern, Switzerland. Available from: https://boris.unibe.ch/id/eprint/178618

40. Horne Z, Powell D, Hummel JE, Holyoak KJ. Countering antivaccination attitudes. Proc Natl Acad Sci. 2015 Aug 18;112(33):10321–4.

41. Schaffer DeRoo S, Pudalov NJ, Fu LY. Planning for a COVID-19 Vaccination Program. JAMA. 2020 Jun 23;323(24):2458.

