## Supplementary material for "Socio-demographic characteristics associated with COVID-19 vaccination uptake in Switzerland: longitudinal analysis of the CoMix study": Supplement_Vaccine_CH_CoMix.pdf

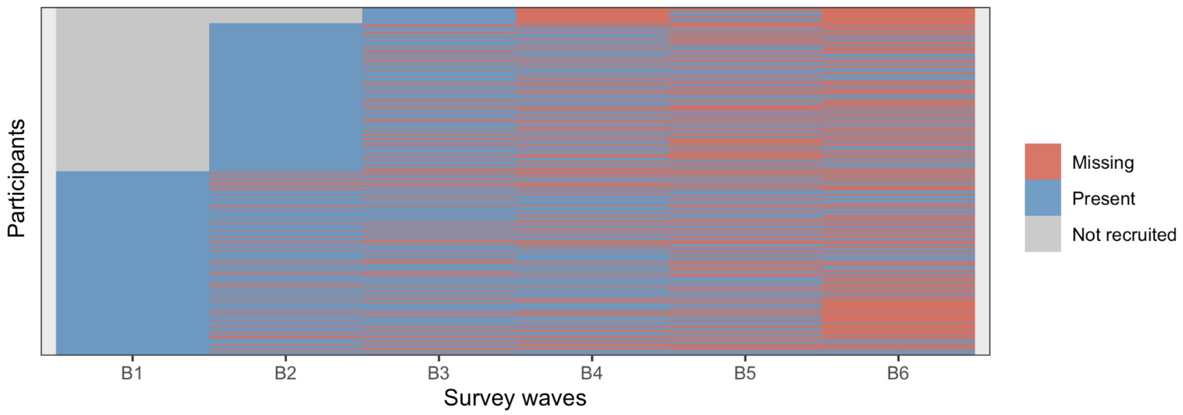


**Supplementary Figure 1:** Participants in the six survey waves of the CoMix study in Switzerland.

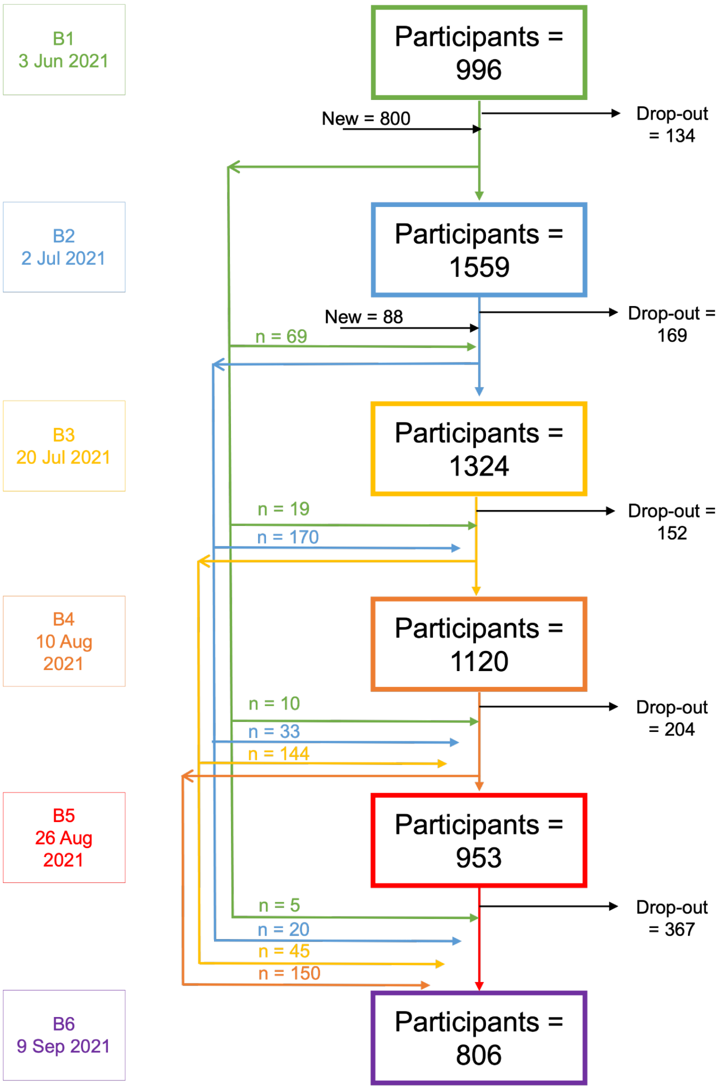


**Supplementary Figure 2**: Flow chart of the participants in the six survey waves of the CoMix study in Switzerland.

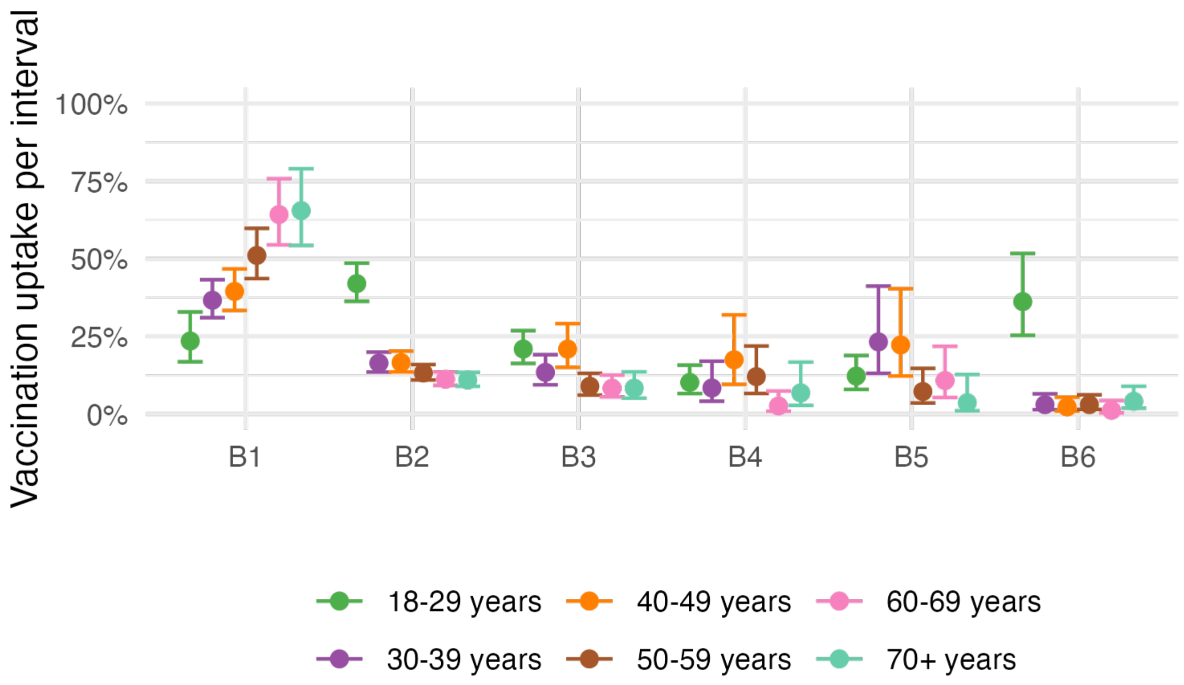


**Supplementary Figure 3:** Vaccination uptake per interval by age group. Vaccination uptake corresponds to the percentage receiving the first vaccine dose amongst those who have not already received it. The adjusted estimates were adjusted for age with an interaction with survey wave, gender, region, Swiss region of residence, country of birth, education level, employment level, net household income, household size, household with a medically vulnerable individual, testing for SARS-CoV-2, number of contacts per day, and attitude towards COVID-19 measures. The error bars indicate the 95% confidence interval.

**Supplementary Table 1:** Milestones of the vaccination program in Switzerland. * Varied across cantons.

| Dates | Description (eligible subpopulation) | **Reference** accessed on 17 May 2023, in German |
|---|---|---|
| 19 Dec 2020 | Swissmedic, the Swiss agency for the authorisation and supervision of therapeutic products, authorised the mRNA vaccine from Pfizer/BioNTech. | https://www.bag.admin.ch/bag/de/home/das-bag/aktuell/medienmitteilungen.msg-id-81667.html |
| 23 Dec 2020 | First person officially received the first vaccine dose in Switzerland. | https://www.srf.ch/news/schweiz/impfstart-in-der-schweiz-luzernerin-erhaelt-erste-impfung-auch-weitere-kantone-gestartet |
| 24 Dec 2020 | Switzerland started the COVID-19 vaccination campaign. Priority was given to the elderly (>75 and then 65-75 years) and the chronically ill, and, secondly*, to healthcare workers and those living with people at risk. | https://www.bag.admin.ch/bag/de/home/das-bag/aktuell/medienmitteilungen.msg-id-81798.html |
| 12 Jan 2021 | Swissmedic authorised the mRNA vaccine from Moderna. | https://www.bag.admin.ch/bag/de/home/das-bag/aktuell/medienmitteilungen.msg-id-81926.html |
| May 2021* | General population (≥16 years) was able to get vaccinated. | |
| 31 Mai 2021 | Vaccinated people were exempt from quarantine, including quarantine after returning from abroad. | https://www.bag.admin.ch/bag/de/home/das-bag/aktuell/medienmitteilungen.msg-id-83531.html |
| June 2021 | Children (≥12 years) were able to get vaccinated. | https://www.bag.admin.ch/bag/de/home/das-bag/aktuell/medienmitteilungen.msg-id-84095.html |
| 13 Sep 2021 | COVID-19 certificate introduced in September 2021: proof of vaccination, recovery or a negative test result were declared mandatory to access indoor hospitality venues, cultural, sporting and leisure activities indoors, and large-scale outdoor events. | https://www.bag.admin.ch/bag/de/home/das-bag/aktuell/medienmitteilungen.msg-id-85035.html |
| 1 Jan 2022 | Children (≥5 years) were able to get vaccinated. | https://www.bag.admin.ch/bag/de/home/das-bag/aktuell/medienmitteilungen.msg-id-86451.html |

**Supplementary Table 2:** Comparison of key characteristics of study participants with the Swiss population. The income of study participants is the household income, whereas the income for the Swiss population corresponds to the individual income from full and part-time employees. Abbreviations: n, number of observations, N, number of participants or population size.

| Category | Variables | Swiss population, N (%) | Study participants, n (%) | Survey wave, N (%) | | | | | |
|---|---|---|---|---|---|---|---|---|---|
| | | | | B1 | B2 | B3 | B4 | B5 | B6 |
| Gender | Female | 4,367,701 (50.4%) | 3,297 (48.8%) | 497 (49.9%) | 761 (48.8%) | 645 (48.7%) | 556 (49.6%) | 452 (47.4%) | 386 (47.9%) |
| | Male | 4,302,599 (49.6%) | 3,436 (50.8%) | 495 (49.7%) | 792 (50.8%) | 673 (50.8%) | 560 (50%) | 498 (52.3%) | 418 (51.9%) |
| | Other | - | 25 (0.4%) | 4 (0.4%) | 6 (0.4%) | 6 (0.5%) | 4 (0.4%) | 3 (0.3%) | 2 (0.2%) |
| Age group | 18-29 | 1,170,597 (16.3%) | 1,044 (15.4%) | 177 (17.8%) | 275 (17.6%) | 207 (15.6%) | 154 (13.8%) | 138 (14.5%) | 93 (11.5%) |
| | 30-39 | 1,239,355 (17.2%) | 1,241 (18.4%) | 180 (18.1%) | 304 (19.5%) | 249 (18.8%) | 202 (18%) | 150 (15.7%) | 156 (19.4%) |
| | 40-49 | 1,200,424 (16.7%) | 1,100 (16.3%) | 159 (16%) | 247 (15.8%) | 219 (16.5%) | 170 (15.2%) | 160 (16.8%) | 145 (18%) |
| | 50-59 | 1,304,794 (18.1%) | 1,407 (20.8%) | 188 (18.9%) | 309 (19.8%) | 273 (20.6%) | 245 (21.9%) | 194 (20.4%) | 198 (24.6%) |
| | 60-69 | 1,005,687 (14%) | 1,159 (17.2%) | 170 (17.1%) | 251 (16.1%) | 222 (16.8%) | 202 (18%) | 179 (18.8%) | 135 (16.7%) |
| | 70+ | 1,276,149 (17.7%) | 807 (11.9%) | 122 (12.2%) | 173 (11.1%) | 154 (11.6%) | 147 (13.1%) | 132 (13.9%) | 79 (9.8%) |
| Household income* | 0-5,000 | 49.4% | 2,150 (31.8%) | 328 (32.9%) | 493 (31.6%) | 425 (32.1%) | 359 (32.1%) | 309 (32.4%) | 236 (29.3%) |
| | 5,001-10,000 | 42.2% | 2,733 (40.4%) | 396 (39.8%) | 634 (40.7%) | 533 (40.3%) | 445 (39.7%) | 384 (40.3%) | 341 (42.3%) |
| | 10,000+ | 7.6% | 861 (12.7%) | 124 (12.4%) | 203 (13%) | 171 (12.9%) | 139 (12.4%) | 122 (12.8%) | 102 (12.7%) |
| | Preferred not to answer | - | 1,014 (15%) | 148 (14.9%) | 229 (14.7%) | 195 (14.7%) | 177 (15.8%) | 138 (14.5%) | 127 (15.8%) |
| Residence | Espace Mittelland | 1,895,693 (21.9%) | 1,489 (22%) | 216 (21.7%) | 344 (22.1%) | 286 (21.6%) | 241 (21.5%) | 226 (23.7%) | 176 (21.8%) |
| | Zurich | 1,553,423 (17.9%) | 1,287 (19%) | 188 (18.9%) | 291 (18.7%) | 244 (18.4%) | 209 (18.7%) | 190 (19.9%) | 165 (20.5%) |
| | Lake Geneva region | 1,669,608 (19.3%) | 1,126 (16.7%) | 168 (16.9%) | 273 (17.5%) | 225 (17%) | 191 (17.1%) | 165 (17.3%) | 104 (12.9%) |
| | Eastern Switzerland | 1,193,069 (13.8%) | 963 (14.2%) | 141 (14.2%) | 219 (14%) | 192 (14.5%) | 161 (14.4%) | 138 (14.5%) | 112 (13.9%) |
| | Northwestern Switzerland | 1,181,776 (13.6%) | 949 (14%) | 143 (14.4%) | 219 (14%) | 186 (14%) | 155 (13.8%) | 132 (13.9%) | 114 (14.1%) |
| | Central Switzerland | 825,745 (9.5%) | 678 (10%) | 96 (9.6%) | 148 (9.5%) | 137 (10.3%) | 108 (9.6%) | 97 (10.2%) | 92 (11.4%) |
| | Ticino | 350,986 (4%) | 266 (3.9%) | 44 (4.4%) | 65 (4.2%) | 54 (4.1%) | 55 (4.9%) | 5 (0.5%) | 43 (5.3%) |

*No absolute values available for net income of the Swiss population.

**Supplementary Table 3:** Comparison of different models to study the association of socio-demographic and other factors with COVID-19 vaccination uptake in Switzerland. Abbreviations: CI, confidence interval; HR, hazard ratio; RR, rate ratio; n, number of observations included in Poisson regression; N, number of participants.

| Categories | Variables | n | N | Cox proportional hazard model | | | | Poisson regression model | | | | |
|---|---|---|---|---|---|---|---|---|---|---|---|---|
| | | | | **Unadjusted HR (95% CI)** | **Unadjusted weighted HR (95% CI)** | **Adjusted HR (95% CI)** | **Adjusted weighted HR (95% CI)** | **Unadjusted RR (95% CI)** | **Unadjusted RR (95% CI) with age as interaction** | **Adjusted RR (95% CI)** | **Adjusted RR (95% CI) with age as interaction** | **Adjusted RR (95% CI) with age as interaction (1 June)** |
| Survey wave<br>Reference: B1 | B2 | 1,143 | - | - | - | - | - | 1.07 (1.01-1.13) | - | - | - | - |
| | B3 | 511 | - | - | - | - | - | 0.43 (0.39-0.48) | - | - | - | - |
| | B4 | 356 | - | - | - | - | - | 0.15 (0.13-0.19) | - | - | - | - |
| | B5 | 277 | - | - | - | - | - | 0.23 (0.19-0.28) | - | - | - | - |
| | B6 | 230 | - | - | - | - | - | 0.20 (0.16-0.25) | - | - | - | - |
| Age groups, years<br>Reference: 18-29 | 30-39 | 737 | 359 | 1.05 (0.87-1.26) | 1.15 (0.98-1.36) | - | - | 1.07 (0.98-1.16) | - | - | - | - |
| | 40-49 | 606 | 314 | 1.09 (0.90-1.32) | 1.15 (0.97-1.36) | - | - | 1.17 (1.07-1.27) | - | - | - | - |
| | 50-59 | 676 | 355 | 1.26 (1.05-1.51) | 1.29 (1.10-1.52) | - | - | 1.30 (1.20-1.42) | - | - | - | - |
| | 60-69 | 474 | 289 | 1.67 (1.39-2.01) | 1.81 (1.51-2.18) | - | - | 1.62 (1.49-1.77) | - | - | - | - |
| | 70+ | 318 | 202 | 2.10 (1.72-2.56) | 2.18 (1.79-2.66) | - | - | 1.85 (1.69-2.03) | - | - | - | - |
| Gender<br>Reference: Female | Male | 1,708 | 955 | 1.17 (1.05-1.30) | 1.05 (0.95-1.16) | 1.07 (0.95-1.20) | 0.96 (0.86-1.07) | 1.17 (1.11-1.23) | 1.12 (1.06-1.17) | 1.09 (1.04-1.15) | 1.09 (1.04-1.15) | 1.11 (1.04-1.19) |
| | Others | 15 | 10 | 1.24 (0.64-2.39) | 0.95 (0.68-1.33) | 0.86 (0.44-1.69) | 0.99 (0.56-1.75) | 1.62 (1.20-2.17) | 1.76 (1.31-2.38) | 1.51 (1.12-2.04) | 1.62 (1.20-2.20) | 1.78 (1.23-2.57) |
| Region<br>Reference: Urban | Rural | 961 | 456 | 0.75 (0.66-0.86) | 0.80 (0.71-0.90) | 0.89 (0.77-1.02) | 0.86 (0.76-0.97) | 0.75 (0.71-0.80) | 0.79 (0.74-0.84) | 0.84 (0.79-0.90) | 0.85 (0.80-0.90) | 0.85 (0.78-0.92) |
| Swiss regions of residence<br>Reference: Espace Mittelland | Zurich | 615 | 351 | 1.22 (1.03-1.44) | 1.14 (0.99-1.33) | 1.23 (1.03-1.47) | 1.11 (0.94-1.30) | 1.23 (1.14-1.33) | 1.19 (1.10-1.29) | 1.10 (1.02-1.20) | 1.11 (1.02-1.20) | 1.12 (1.01-1.25) |
| | Lake Geneva region | 590 | 337 | 1.12 (0.94-1.33) | 1.06 (0.92-1.23) | 1.02 (0.86-1.22) | 1.02 (0.88-1.19) | 1.17 (1.08-1.27) | 1.13 (1.05-1.23) | 1.06 (0.98-1.15) | 1.06 (0.98-1.15) | 1.05 (0.95-1.16) |
| | Eastern Switzerland | 501 | 263 | 1.09 (0.91-1.32) | 1.03 (0.88-1.21) | 1.14 (0.95-1.38) | 1.10 (0.93-1.30) | 1.10 (1.01-1.20) | 1.09 (1.00-1.19) | 1.10 (1.01-1.20) | 1.09 (1.00-1.18) | 1.09 (0.98-1.22) |
| | Northwestern Switzerland | 262 | 262 | 1.11 (0.92-1.34) | 1.07 (0.91-1.26) | 1.28 (1.05-1.55) | 1.17 (0.99-1.39) | 1.07 (0.98-1.16) | 1.08 (0.99-1.17) | 1.08 (0.99-1.18) | 1.07 (0.98-1.17) | 1.09 (0.97-1.22) |
| | Central Switzerland | 352 | 182 | 1.07 (0.87-1.32) | 0.97 (0.80-1.17) | 1.23 (0.99-1.52) | 1.06 (0.86-1.31) | 1.05 (0.95-1.16) | 1.11 (1.01-1.23) | 1.18 (1.07-1.30) | 1.16 (1.05-1.28) | 1.20 (1.06-1.36) |
| | Ticino | 140 | 82 | 1.29 (0.97-1.72) | 1.59 (1.11-2.27) | 1.50 (1.12-2.02) | 1.74 (1.20-2.53) | 1.16 (1.02-1.32) | 1.16 (1.02-1.33) | 1.14 (1.00-1.30) | 1.15 (1.01-1.31) | 1.18 (1.00-1.40) |
| Country of birth<br>Reference: Switzerland | EU | 450 | 249 | 1.10 (0.93-1.29) | 1.06 (0.93-1.22) | 1.09 (0.92-1.29) | 1.09 (0.94-1.27) | 1.05 (0.98-1.13) | 1.02 (0.94-1.10) | 0.97 (0.90-1.04) | 0.96 (0.89-1.04) | 0.97 (0.88-1.07) |
| | Non-EU | 283 | 156 | 1.11 (0.91-1.35) | 1.13 (0.96-1.33) | 1.03 (0.84-1.25) | 1.08 (0.89-1.30) | 1.06 (0.97-1.16) | 1.01 (0.93-1.11) | 0.96 (0.88-1.05) | 0.98 (0.89-1.07) | 1.01 (0.90-1.13) |
| | Unknown | 235 | 147 | 1.18 (0.97-1.45) | 1.11 (0.86-1.44) | 1.10 (0.89-1.36) | 1.07 (0.83-1.38) | 1.18 (1.08-1.29) | 1.03 (0.94-1.13) | 1.06 (0.97-1.16) | 1.06 (0.97-1.17) | 1.10 (0.97-1.24) |
| Education level<br>Reference: Lowest level | Middle level of education | 1,225 | 639 | 1.08 (0.95-1.22) | 1.11 (0.99-1.25) | 1.00 (0.88-1.14) | 1.04 (0.92-1.17) | 1.06 (1.00-1.12) | 1.06 (1.00-1.13) | 1.01 (0.95-1.08) | 1.01 (0.95-1.07) | 0.99 (0.92-1.07) |
| | Highest level of education | 714 | 439 | 1.41 (1.23-1.61) | 1.23 (1.10-1.37) | 1.26 (1.08-1.46) | 1.17 (1.02-1.33) | 1.39 (1.31-1.48) | 1.35 (1.27-1.43) | 1.18 (1.10-1.26) | 1.18 (1.10-1.27) | 1.19 (1.09-1.30) |
| Employment status<br>Reference: Employed | Unemployed | 214 | 110 | 0.88 (0.69-1.13) | 1.04 (0.77-1.40) | 0.89 (0.69-1.15) | 1.04 (0.77-1.42) | 0.86 (0.77-0.96) | 0.83 (0.74-0.93) | 0.86 (0.77-0.97) | 0.86 (0.76-0.97) | 0.88 (0.76-1.03) |
| | Student | 192 | 116 | 1.23 (0.98-1.54) | 1.04 (0.86-1.25) | 1.55 (1.17-2.05) | 1.27 (0.98-1.64) | 1.13 (1.02-1.26) | 1.38 (1.22-1.56) | 1.26 (1.11-1.43) | 1.33 (1.17-1.51) | 1.35 (1.15-1.58) |
| | Homemaker | 164 | 75 | 0.73 (0.54-0.99) | 0.75 (0.58-0.97) | 0.95 (0.69-1.32) | 0.81 (0.63-1.05) | 0.77 (0.67-0.89) | 0.80 (0.70-0.92) | 0.96 (0.83-1.10) | 0.95 (0.82-1.10) | 0.97 (0.81-1.16) |
| | Retired | 607 | 377 | 1.66 (1.45-1.89) | 1.63 (1.44-1.85) | 1.20 (0.95-1.51) | 1.11 (0.89-1.38) | 1.46 (1.38-1.55) | 1.01 (0.91-1.12) | 1.05 (0.95-1.17) | 1.05 (0.94-1.16) | 1.08 (0.94-1.24) |
| | Other unemployed situation | 92 | 44 | 0.85 (0.58-1.26) | 1.07 (0.78-1.47) | 0.96 (0.64-1.44) | 1.30 (0.91-1.85) | 0.85 (0.71-1.01) | 0.82 (0.69-0.98) | 0.92 (0.77-1.10) | 0.90 (0.75-1.07) | 0.80 (0.63-1.03) |
| Household income, net | 5,001-10,000 CHF | 1,403 | 762 | 1.19 (1.04-1.36) | 1.14 (1.01-1.28) | 1.25 (1.08-1.44) | 1.24 (1.08-1.42) | 1.18 (1.11-1.25) | 1.21 (1.14-1.28) | 1.15 (1.08-1.23) | 1.15 (1.08-1.23) | 1.14 (1.04-1.24) |

| Categories | Variables | n | N | Cox proportional hazard model | | | | Poisson regression model | | | | |
|---|---|---|---|---|---|---|---|---|---|---|---|---|
| Reference: 0-5,000 CHF | | | | | | | | | | | | |
| | 10,000+ CHF | 387 | 248 | 1.49 (1.26-1.77) | 1.25 (1.08-1.45) | 1.42 (1.17-1.73) | 1.29 (1.09-1.54) | 1.50 (1.39-1.62) | 1.47 (1.36-1.59) | 1.33 (1.21-1.45) | 1.34 (1.23-1.46) | 1.36 (1.22-1.53) |
| | Preferred not to answer | 529 | 281 | 1.16 (0.98-1.38) | 1.17 (1.00-1.37) | 1.14 (0.95-1.36) | 1.15 (0.98-1.36) | 1.17 (1.08-1.27) | 1.16 (1.07-1.25) | 1.14 (1.06-1.24) | 1.13 (1.04-1.23) | 1.15 (1.03-1.27) |
| Household size | Mean (range) | 2 (1 - 10) | 2 (1 - 10) | 0.96 (0.92-1.00) | 0.95 (0.91-0.98) | 0.95 (0.89-1.01) | 0.97 (0.92-1.02) | 0.97 (0.95-0.98) | 1.02 (0.99-1.04) | 0.96 (0.94-0.99) | 0.96 (0.94-0.99) | 0.97 (0.94-1.01) |
| Household with medically vulnerability<br>Reference: No person in a risk group | One or more person in a risk group | 944 | 578 | 1.33 (1.19-1.49) | 1.23 (1.11-1.37) | 1.21 (1.07-1.36) | 1.18 (1.05-1.33) | 1.36 (1.30-1.44) | 1.20 (1.13-1.26) | 1.18 (1.11-1.24) | 1.16 (1.10-1.23) | 1.16 (1.08-1.25) |
| Testing for SARS-CoV-2<br>Reference: Tested positive | Tested | 872 | - | 0.85 (0.46-1.55) | 1.11 (0.73-1.70) | 0.98 (0.53-1.82) | 1.23 (0.77-1.95) | 0.73 (0.55-0.98) | 0.75 (0.56-1.00) | 0.85 (0.63-1.13) | 0.87 (0.65-1.17) | 0.76 (0.55-1.06) |
| | Never tested | 2,557 | - | 1.72 (0.95-3.11) | 1.72 (1.14-2.58) | 1.96 (1.07-3.59) | 1.84 (1.17-2.90) | 1.09 (0.82-1.44) | 0.87 (0.65-1.15) | 1.02 (0.77-1.37) | 1.06 (0.80-1.42) | 0.96 (0.69-1.32) |
| | Preferred not to answer | 56 | - | 0.96 (0.43-2.15) | 1.13 (0.55-2.31) | 1.59 (0.70-3.61) | 1.53 (0.70-3.35) | 0.58 (0.40-0.85) | 0.55 (0.38-0.81) | 0.74 (0.50-1.09) | 0.76 (0.51-1.12) | 0.61 (0.38-0.98) |
| Number of contacts per day<br>Reference: 0-2 | 3-5 | 937 | - | 1.13 (0.98-1.30) | 1.08 (0.95-1.22) | 1.23 (1.07-1.43) | 1.13 (0.99-1.29) | 1.03 (0.97-1.09) | 1.02 (0.96-1.09) | 1.02 (0.95-1.09) | 1.02 (0.95-1.09) | 1.01 (0.92-1.10) |
| | 6+ | 1,282 | - | 1.16 (1.02-1.31) | 1.01 (0.90-1.13) | 1.28 (1.10-1.49) | 1.13 (0.98-1.30) | 1.06 (1.00-1.12) | 1.09 (1.03-1.16) | 1.08 (1.01-1.16) | 1.08 (1.01-1.16) | 1.10 (1.01-1.20) |
| Attitudes towards COVID-19 measures<br>Reference: About right | Too lenient | 663 | - | 0.88 (0.77-1.01) | 0.81 (0.73-0.90) | 0.85 (0.74-0.97) | 0.79 (0.70-0.88) | 1.02 (0.96-1.08) | 1.05 (0.99-1.12) | 1.01 (0.95-1.07) | 1.02 (0.96-1.08) | 1.03 (0.95-1.11) |
| | Too strict | 1,157 | - | 0.32 (0.28-0.37) | 0.49 (0.41-0.58) | 0.33 (0.28-0.39) | 0.51 (0.42-0.60) | 0.44 (0.41-0.47) | 0.55 (0.52-0.59) | 0.56 (0.52-0.60) | 0.56 (0.53-0.61) | 0.52 (0.48-0.57) |
| | Don't know | 111 | - | 0.43 (0.29-0.63) | 0.53 (0.38-0.74) | 0.45 (0.31-0.68) | 0.56 (0.39-0.80) | 0.51 (0.43-0.61) | 0.61 (0.51-0.72) | 0.63 (0.53-0.76) | 0.64 (0.53-0.77) | 0.70 (0.56-0.88) |

**Supplementary Table 4:** Cox proportional hazard model to study the association of socio-demographic and other factors with COVID-19 vaccination uptake in Switzerland. Compared to Supplementary Table 3, Supplementary Table 4 shows participants that either got not vaccinated during our study or had an exact date of vaccination. Abbreviations: CI, confidence interval; HR, hazard ratio; N, number of participants.

| Categories | Variables | N | Unadjusted HR (95% CI) | Adjusted weighted HR (95% CI) |
|---|---|---|---|---|
| Age groups, years<br>Reference: 18-29 | 30-39 | 336 | 1.31 (1.07-1.61) | |
| | 40-49 | 282 | 1.31 (1.06-1.62) | |
| | 50-59 | 334 | 1.89 (1.55-2.31) | |
| | 60-69 | 266 | 2.98 (2.42-3.66) | |
| | 70+ | 186 | 4.78 (3.83-5.97) | |
| Gender<br>Reference: Female | Male | 869 | 1.05 (0.93-1.17) | 0.95 (0.84-1.07) |
| | Others | 7 | 2.29 (0.95-5.53) | 2.23 (0.90-5.56) |
| Region<br>Reference: Urban | Rural | 430 | 0.89 (0.78-1.03) | 0.98 (0.85-1.15) |
| Swiss regions of residence<br>Reference: Espace Mittelland | Zurich | 323 | 1.12 (0.93-1.34) | 0.98 (0.82-1.18) |
| | Lake Geneva region | 316 | 1.12 (0.93-1.35) | 1.09 (0.90-1.32) |
| | Eastern Switzerland | 247 | 1.10 (0.90-1.34) | 1.05 (0.86-1.27) |
| | Northwestern Switzerland | 231 | 1.08 (0.87-1.33) | 1.13 (0.91-1.39) |
| | Central Switzerland | 161 | 0.87 (0.69-1.09) | 0.95 (0.76-1.20) |
| | Ticino | 77 | 1.01 (0.74-1.36) | 1.24 (0.83-1.85) |
| Country of birth<br>Reference: Switzerland | EU | 237 | 0.99 (0.84-1.17) | 0.95 (0.80-1.12) |
| | Non-EU | 150 | 0.95 (0.77-1.17) | 0.93 (0.76-1.14) |
| | Unknown | 117 | 1.10 (0.87-1.40) | 1.08 (0.82-1.41) |
| Education level<br>Reference: Lowest level | Middle level of education | 584 | 1.01 (0.88-1.16) | 1.17 (1.01-1.35) |
| | Highest level of education | 397 | 1.02 (0.89-1.19) | 1.26 (1.08-1.47) |
| Employment status<br>Reference: Employed | Unemployed | 104 | 1.01 (0.77-1.31) | 1.03 (0.76-1.39) |
| | Student | 97 | 0.71 (0.55-0.92) | 0.95 (0.70-1.27) |
| | Homemaker | 68 | 0.99 (0.70-1.40) | 1.00 (0.72-1.38) |
| | Retired | 348 | 2.47 (2.15-2.85) | 1.24 (0.98-1.57) |
| | Other unemployed situation | 41 | 1.83 (1.21-2.78) | 1.56 (1.06-2.29) |
| Household income, net<br>Reference: 0-5,000 CHF | 5,001-10,000 CHF | 704 | 0.95 (0.83-1.10) | 1.36 (1.16-1.60) |
| | 10,000+ CHF | 231 | 0.90 (0.75-1.08) | 1.26 (1.01-1.56) |
| | Preferred not to answer | 238 | 0.91 (0.75-1.11) | 1.02 (0.84-1.24) |
| Household size | Mean (range) | 2 (1 - 9) | 0.85 (0.81-0.90) | 0.88 (0.82-0.94) |
| Household with medically vulnerability<br>Reference: No person in a risk group | One or more person in a risk group | 529 | 1.46 (1.29-1.65) | 1.50 (1.31-1.72) |
| Testing for SARS-CoV-2<br>Reference: Tested positive | Tested | 316 | 0.81 (0.40-1.63) | 0.75 (0.33-1.73) |
| | Never Tested | 1376 | 1.37 (0.68-2.75) | 1.06 (0.46-2.41) |
| | Preferred not to answer | 25 | 0.56 (0.21-1.49) | 0.56 (0.19-1.64) |
| Number of contacts per day<br>Reference: 0-2 | 3-5 | 462 | 1.01 (0.87-1.17) | 1.17 (1.01-1.37) |
| | 6+ | 692 | 0.87 (0.76-0.99) | 1.11 (0.94-1.30) |
| Attitudes towards COVID-19 measures<br>Reference: About right | Too lenient | 368 | 0.94 (0.82-1.08) | 0.84 (0.73-0.96) |
| | Too strict | 420 | 0.74 (0.62-0.87) | 0.82 (0.69-0.97) |
| | Don't know | 46 | 0.39 (0.24-0.62) | 0.72 (0.36-1.44) |

**Supplementary Table 5:** Results from the logistic regression model to study the association of study participants' characteristics with missed survey waves. Abbreviations: CI, confidence interval; OR, odds ratio; aOR, adjusted OR; N, number of participants.

| **Categories** | **Variables** | **N** | **Univariable OR (95% CI)** | **aOR (95% CI)** without time varying variables | **aOR (95% CI)** with time varying variables |
|---|---|---|---|---|---|
| Age groups, years<br>Reference: 18-29 | 30-39 | 358 | 0.54 (0.36-0.81) | 0.94 (0.88-1.00) | 0.93 (0.87-1.00) |
| | 40-49 | 308 | 0.41 (0.27-0.61) | 0.90 (0.83-0.96) | 0.89 (0.83-0.95) |
| | 50-59 | 363 | 0.28 (0.19-0.41) | 0.84 (0.78-0.90) | 0.84 (0.78-0.90) |
| | 60-69 | 289 | 0.35 (0.23-0.52) | 0.90 (0.82-0.98) | 0.90 (0.82-0.98) |
| | 70+ | 207 | 0.37 (0.24-0.57) | 0.91 (0.82-1.02) | 0.92 (0.82-1.03) |
| Gender<br>Reference: Female | Male | 955 | 0.91 (0.74-1.13) | 0.98 (0.94-1.02) | 0.98 (0.95-1.02) |
| | Others | 10 | 1.17 (0.25-5.56) | 1.03 (0.79-1.34) | 1.04 (0.80-1.34) |
| Region<br>Reference: Urban | Rural | 457 | 0.86 (0.67-1.09) | 0.99 (0.94-1.04) | 0.99 (0.94-1.03) |
| Swiss region of residence<br>Reference: Espace Mittelland | Zurich | 351 | 0.97 (0.70-1.34) | 0.99 (0.93-1.05) | 0.99 (0.93-1.05) |
| | Lake Geneva region | 337 | 1.66 (1.16-2.38) | 1.08 (1.01-1.14) | 1.07 (1.01-1.13) |
| | Eastern Switzerland | 263 | 1.08 (0.75-1.54) | 1.01 (0.94-1.07) | 1.01 (0.94-1.07) |
| | Northwestern Switzerland | 262 | 1.16 (0.81-1.67) | 1.02 (0.95-1.09) | 1.02 (0.95-1.09) |
| | Central Switzerland | 182 | 0.88 (0.60-1.31) | 0.98 (0.91-1.06) | 0.99 (0.92-1.06) |
| | Ticino | 82 | 5.44 (2.14-13.81) | 1.22 (1.10-1.35) | 1.21 (1.10-1.34) |
| Country of birth<br>Reference: Switzerland | EU | 249 | 1.61 (1.13-2.30) | 1.09 (1.03-1.16) | 1.10 (1.03-1.16) |
| | Non-EU | 156 | 1.32 (0.87-1.99) | 1.05 (0.98-1.12) | 1.05 (0.98-1.12) |
| | Unknown | 147 | 0.85 (0.58-1.24) | 0.99 (0.92-1.06) | 0.98 (0.91-1.06) |
| Education level<br>Reference: Lowest level | Middle level of education | 639 | 1.03 (0.81-1.31) | 0.98 (0.94-1.03) | 0.97 (0.93-1.02) |
| | Highest level of education | 439 | 1.18 (0.89-1.56) | 0.99 (0.93-1.04) | 0.99 (0.94-1.04) |
| Employment status<br>Reference: Employed | Unemployed | 110 | 1.05 (0.66-1.67) | 1.01 (0.92-1.09) | 1.00 (0.92-1.09) |
| | Student | 116 | 5.64 (2.45-12.96) | 1.08 (0.99-1.19) | 1.09 (0.99-1.20) |
| | Homemaker | 75 | 0.70 (0.42-1.16) | 0.92 (0.83-1.02) | 0.92 (0.83-1.02) |
| | Retired | 377 | 0.83 (0.64-1.08) | 1.00 (0.92-1.08) | 1.00 (0.92-1.09) |
| | Other unemployed situation | 44 | 0.73 (0.38-1.42) | 0.98 (0.86-1.12) | 0.97 (0.86-1.11) |
| Household income, net<br>Reference: 0-5,000 CHF | 5,001-10,000 CHF | 762 | 0.98 (0.76-1.27) | 0.98 (0.93-1.03) | 0.98 (0.93-1.02) |
| | 10,000+ CHF | 248 | 1.07 (0.75-1.52) | 0.98 (0.91-1.05) | 0.99 (0.93-1.06) |
| | Preferred not to answer | 281 | 0.93 (0.67-1.30) | 0.97 (0.91-1.03) | 0.98 (0.92-1.04) |
| Household size | Mean (range) | 2 (1 - 10) | 1.20 (1.09-1.32) | 1.02 (1.01-1.04) | 0.99 (0.98-1.01) |
| Household with medically vulnerability<br>Reference: No person in a risk group | One or more person in a risk group | 578 | 0.84 (0.67-1.06) | 0.97 (0.93-1.02) | 0.97 (0.93-1.02) |
| Vaccination status<br>Reference: Not vaccinated | Vaccinated | 1,321 | 0.74 (0.58-0.94) | - | 0.95 (0.91-0.99) |
| Testing for SARS-CoV-2<br>Reference: Tested positive | Tested | 543 | 0.66 (0.25-1.76) | - | 0.94 (0.81-1.09) |
| | Never tested | 1,277 | 0.60 (0.23-1.58) | - | 0.95 (0.81-1.10) |
| | Preferred not to answer | 32 | 0.83 (0.23-3.07) | - | 0.98 (0.79-1.21) |
| Number of contacts per day<br>Reference: 0-2 | 3-5 | 527 | 1.12 (0.87-1.43) | - | 1.02 (0.98-1.07) |
| | 6+ | 589 | 2.54 (1.92-3.36) | - | 1.16 (1.10-1.23) |
| Attitudes towards COVID-19 measures<br>Reference: About right | Too lenient | 423 | 1.07 (0.81-1.41) | - | 1.03 (0.98-1.08) |
| | Too strict | 501 | 0.90 (0.70-1.16) | - | 0.97 (0.92-1.01) |
| | Don't know | 46 | 1.71 (0.75-3.87) | - | 1.08 (0.94-1.23) |
